# Geographical Distance to the Epicenter of Covid-19 Predicts the Burnout of Working Population: Ripple effect or Typhoon Eye effect?

**DOI:** 10.1101/2020.04.02.20051128

**Authors:** Stephen X Zhang, Hao Huang, Feng Wei

**Affiliations:** University of Adelaide, Adelaide, Australia; Southwestern University of Finance and Economics, Chengdu, China; Tongji University, Shanghai, China

**Keywords:** burnout, distance to the epicenter, ripple effect, typhoon eye effect, epidemic, pandemic, coronavirus, SARS-CoV-2

## Abstract

COVID-19 originated in Wuhan and rippled across China. We investigate how the geographical distance of working adults to the epicenter of Wuhan predicts their burnout. Preliminary results of a survey of 308 working adults in 53 cities showed working adults’ distance to the epicenter of Wuhan had an inverted U-shaped relationship with their burnout. Such results help to identify regions where people would need more psychiatric assistance, carrying direct implications to healthcare practitioners and policymakers.

## 1. Introduction

Covid-19 set off in Wuhan in December 2019. Wuhan, a metropolitan with over 11 million population, is a key transportation hub in central China. The movement of the people helped the virus to steadily sweep across China by the end of Jan 2020. By Feb 2020, almost all people in China had their lives and work disrupted (Zhang et al 2020), running the risking of burnout – an emotional, physical, and mental exhaustion due to excessive and prolonged stress of being overwhelmed, emotionally drained, and incapable to meet constant demands (Maslach & Leiter, 2016). Who may be most susceptible to burnout during this crisis?

Early research suggested the impact of a crisis spreads out in a circle and hence declines gradually over geographical distance, known as the ripple effect (Slovic, 1987). However, evidence from the fields often found the opposite – the level of anxiety and concerns was lower for residents who were nearer to a nuclear power plant (Guedeney and Mendel, 1973), an epicenter of an earthquake (Li et al., 2009), a risky mine (Zheng et al., 2015), as well the epicenter of SARS (Shi et al., 2003). Such an effect is termed a typhoon eye effect, as the epicenter of a typhoon is relatively calm.

The theories of “ripple effect” and “psychological typhoon eye effect” offer opposite predictions, and we aim to examine these opposing predictions by analyzing the relationship between working adults’ distance to the epicenter of Covid-19 and their burnout. Burnout is typically associated with cardiovascular problems, headaches, chronic fatigue, gastrointestinal disorders, muscle tension, hypertension, cold/flu episodes, and sleep disturbances, and burnout in working population predicts job dissatisfaction, negative impacts on their colleagues, lower productivity and impaired quality of work, low organizational commitment, absenteeism, and turnover (Maslach & Leiter, 2016).

As the world now is weathering the storm of Covid-19, this study helps psychotherapists and healthcare policy-maker to identify those in more susceptible areas for prioritizing assistance during the Covid-19 outbreak (Liu et al., 2020).

## 2. Method

Covid-19 began in late 2019 at Wuhan, and Covid-19 came into the public eyes on Jan 20, 2020 when the Chinese premier publicly urged decisive and effective efforts to prevent and control the virus. Two days later, on 22 Jan, China decided to shut down Wuhan, stopping all modes of transportation by the morning of the next day. However, on the night of the 22nd, about 300 000 people left by train alone. About 70% of the people left Wuhan for other locations within the same province; 14% went to the neighboring provinces of Henan, Hunan, Anhui and Jiangxi; and the rest went further (Kinetz, 2020). The virus kept on spreading across China, and we conducted a survey on February 20–21, 2020 on working adults from locations that vary in their travelling distance from Wuhan. We reached 410 adults, and 308 of them answered the survey, with a response rate of 75.1%. All respondents agreed to participate in the study, which was approved by the ethics committee at Tongji University (#20200211).

The participants provided their socio-demographic characteristics, such as gender, age, education, job status (worked at office, worked from home, or suspended working), and their locations at the time of the survey. Using their locations, we calculated each participant’s distance to Wuhan. We assessed burnout over the previous month using the 5-item Chinese version of Maslach Burnout Inventory-General Survey (MBI-GS). We used Stata 16 for OLS regression analysis to analyze the relationship between the distance to the epicenter and burnout.

## 3. Results

The regression analysis uncovers an inverted U-shaped relationship between working adults’ distance to the epicenter and their burnout (β=-0.275, p=0.036) (see Table 1). The inflection point of the inverted U-shaped relationship lies at 1020 km from the epicenter. Such an inverted U-shaped relationship suggests both ripple effect and typhoon eye effect to take place, where typhoon eye effect dominated close to the epicenter and ripple effect dominated further away.

**Table 1.** Working adults’ burnout by their distance to the epicenter

|  | Burnout |  |  |
| --- | --- | --- | --- |
|  | Coefficient | SE | 95% CI |
| **Covariates** |  |  |  |
| Gender | -0.053 | 0.075 | [-0.009, 1.130] |
| Age | -0.007 | 0.004 | [-0.016, 0.007] |
| Education | 0.090* | 0.031 | [0.028, 0.151] |
| Worked at office (reference group) |  |  |  |
| Worked from home | 0.046 | 0.090 | [-0.133, 0.225] |
| Stopped working | 0.048 | 0.102 | [-0.154, 0.250] |
| **Independent Variable** |  |  |  |
| Distance to the epicenter | 0.561 | 0.289 | [-0.008, 1.130] |
| **Quadratic Effect** |  |  |  |
| Distance to the epicenter squared | -0.275* | 0.131 | [-0.532, -0.018] |

N=308. \* $p<0.05$ , \*\* $p<0.01$ , \*\*\* $p<0.001$ .

## 4. Discussions

Unlike prior studies that examined either ripple effect or typhoon eye effect from the epicenter of crises, we found an inverted U-shaped relationship, suggesting the two effects both in play. Our results suggest psychiatrists and public health policymakers need to be careful using either theory alone in identifying and screening people at mental health risk based on geographical distance.

We note our limitations that offer opportunities for future research. The inversed U-shaped relationship that we found was limited to predicting burnout, a psychological syndrome under prolonged social stressors, and it is interesting to examine how the distance to epicenter predicts other mental health outcomes. The inversed U-shaped relationship is also limited to China, which had one clear epicenter in Wuhan, where Covid-19 originated. China is also a geographically large country, where the epicenter of Wuhan happens to be centrally located. Hence, ripple effect and typhoon eye effect may play out differently in other countries with multiple epicenters of Covid-19 (e.g. Washington State and New York State in the US) and distinct geographical features.

Nonetheless, our results point to the need for further studies to determine how the two effects may dominate each other in other Covid-19 infected areas to enable better identification of those who are in greater need.

## Data Availability

Data are available upon request

## Acknowledgement

The data collection was funded by the Chinese National Funding of Social Sciences (Grant number: 1509093).

